# Using Whole Genome Sequencing to Characterize Clinically Significant Blood Groups Among Healthy Older Australians

**DOI:** 10.1101/2021.04.18.21255241

**Authors:** Sudhir Jadhao, Candice Davison, Eileen V. Roulis, Simon Lee, Paul Lacaze, Moeen Riaz, John J McNeil, David M Thomas, Natalie M. Pecheniuk, Catherine A. Hyland, Robert L. Flower, Shivashankar H. Nagaraj

## Abstract

There have been no comprehensive studies of a full range of blood group polymorphisms within the Australian population. The problem is compounded by the absence of any databases carrying genomic information on chronically transfused patients and low frequency blood group antigens in Australia. Here, we use RBCeq, a web server-based blood group genotyping software, to identify unique blood group variants among Australians and compare the variation detected versus global data. Whole genome sequencing data was analysed from for 2796 healthy older Australians from the Medical Genome Reference Bank and compared with data from 1000G phase 3 (1KGP3) databases comprising 661 African, 347 American, 503 European, 504 East Asian, and 489 South Asian participants. There were 688 rare variants detected in this Australian sample population, including nine variants that had clinical associations. Notably, we identified 149 variants that were computationally predicted to be novel and deleterious. No clinically significant rare or novel variants were found associated with the genetically complex ABO blood group system. For the Rh blood group system, one novel and 16 rare variants were found. Our detailed blood group profiling results provide a starting point for the creation of an Australian blood group variant database.

**Key points:**

- We identified unique blood group variants among the healthy older Australian population compared with global data using RBCeq software.
- Our detailed blood group profiling result may be a starting point for the creation of an Australian blood group variant database.

## INTRODUCTION

Modern transfusion medicine has highlighted the geographical and ethnic variability in blood group allele and genotype frequencies. The International Society of Blood Transfusion (ISBT) has recognised 43 red blood cell RBC membrane-associated blood group systems, involving over 360 antigen phenotypes defined by >1500 alleles across 45 genes and two transcription factors (TF).^1-4^ Blood group genes encode multiple structurally and functionally distinct molecules and exhibit varying degrees of polymorphic complexity (insertions/deletions [indels], single nucleotide variants [SNVs], copy number variations [CNVs], and structural variants [SVs]) within the population.^5,6^ Misidentification of any of these variants may contribute towards alloimmunizationof individuals requiring RBC transfusion support, which introduces a risk of adverse events such as Hemolytic Transfusion Reactions (HTRs), Hemolytic Disease of the Fetus and Newborn (HDFN), pregnancy complications, or more subtle allergic reactions of clinical significance ^7^.

Routinely in Australia, the Australian New Zealand Society of Blood Transfusion (ANZSBT) guidelines for Transfusion and Immunohematology laboratory practice advise to select ABO and RhD matched red cell products for transfusion to ensure transfusion safety.^8,9 10^ The RH (e.g., D, C/c, E/e) and MNS (e.g., M/N, S/s, U) blood group antigens are encoded by multiple complex alleles, genetic variations, and gene rearrangements between *RHD/RHCE* and *GYPA/GYPB/GYPE* with distinct population specific distributions. For example, the D negative blood type is prevalent in groups with Caucasian (15%)^11^ and African (8%) ancestries but rare amongst Asians (<0.1%)^12^. The variations in blood group antigen expression also may increase or decrease host susceptibility to infections in some diseases. The glycoprotein that carries the antigens of the Duffy blood group system (FY) is a typical chemokine receptor 1 (ACKR1), also previously known as Duffy Antigen Receptor for Chemokines (DARC). ACKR1 acts as a receptor for *Plasmodium vivax* and *Plasmodium knowlesi* malaria parasites. The absence of FY glycoprotein genotype is common among Africans and African-Americans, conferring these populations with resistance to malaria infection.^13,14^ Variations in *ACKR1* are also associated with a survival advantage in leukopenic HIV patients.^15^ The recessive African-specific *ACKR1* null allele increases the risk of HIV-1 infection.^14^ In the ABO blood group system, individuals with group O blood type are more prone to vaso-occlusive crises associated with sickle cell disease (SCD).^16^ However, the sickle hemoglobin (HbS) of SCD combined with O-type blood group confers some resistance to *Plasmodium falciparum* infection.^17^ O-blood type has also been found to confer resistance towards developing cardiovascular disease or type-2 diabetes.

Blood group typing by traditional serological, molecular or SNP microarray methods have limited functionality in characterizing blood group antigens which are rare, have weak expression or recombinant, partial, or novel.^18^ There is mounting evidence that clinically significant rare antigens and novel variants confound conventional serologic typing and SNP approaches. ^19^ To achieve extended RBC typing, a more comprehensive approach would be to apply Next-Generation Sequencing (NGS) to overcome the limitations of serological and molecular techniques.^20-24^ NGS approaches including targeted exome sequencing (TES), whole exome sequencing (WES), and whole genome sequencing (WGS) have the potential to provide a new basis of pre-transfusion testing by facilitating the accurate characterization of an individual’s complete blood group profile, supporting precision-based medicine.^25,26^ Curated and detailed DNA-phenotype-annotated databases storing blood group allele and antigen data has been developed by ISBT, providing tools for blood type calling from genetic data. As NGS is increasingly employed in immunohematology, Erythrogene, a custom-designed blood group allele database associated with the 1000 Genomes (1KGP3) project has been developed and applied.^6,27^

Accurate prediction of blood group phenotypes based on NGS data requires immuno-genetic knowledge since multiple genotypes can lead to the same phenotype (e.g. ABO, MNS and LE systems) and not all blood group antigens are direct products of primary genes like the ABO, LE and H systems.^25^ Even though various tools/algorithms have been developed applying statistical and machine learning approaches to process NGS data,^28,29^ there is no single tool which provides complete and comprehensive automation of blood group characterization in user-friendly manner. Only two tools are available to date, BOOGIE^30^ and BloodTyper,^25^ neither of which has the potential to identify novel blood group variants. To overcome this limitation, we developed a comprehensive and secure bioinformatics platform called RBCeq. RBCeq (https://www.rbceq.org/) is a web server-based blood group genotyping software. It is able to provide fast and accurate extended mass screening of blood groups, including from diverse populations with distinctive and complex blood group profiles.

Blood group antigen profiles are well characterized for European, North American, and some East Asian populations, but no extensive study has been carried out to date on the blood group antigen profile of the Australian population. Australia constitutes a highly heterogeneous multi-cultural and multi-ethnic population. ^31 32^ The patient population in Australia is more diverse than blood donors and there is a greater need in patients for blood types that are rare in a Caucasian population. Most donors are Caucasian, previous studies have indicated that immigrants and ethnic minorities are less involved in blood donation^33^. To date, there has not been any study on the blood group polymorphisms within the Caucasian Australian population as a whole. The problem is compounded by the absence of rare blood group antigen reports on a large number of Australian individuals. However, recently the Medical Genome Reference Bank (MGRB) was released comprising WGS data of 4000 healthy elderly Australians mostly of European descent.^34,35^ These individuals are participants of the ASPirin in Reducing Events in the Elderly (ASPREE) study, initiated to investigate whether the daily use of aspirin would prolong the healthy life span of older adults. The largest ongoing study of healthy ageing in the Southern Hemisphere^36^. In this study, we use genomic data from the MGRB ASPREE participants to identify unique blood group variants among a healthy older Australian general population and compared this with global data. To the best of our knowledge, this is the first study using WGS data for identifying the prevalence of blood group polymorphisms within a large Australian population dataset compared with global trends. Results of this study will help characterize key blood group systems and the frequency of rare phenotypes amongst the Australian general population, providing a tool to improve treatment accuracy and reduce the rate of hemolytic transfusion reactions. Also, providing an important proof-of-concept for using WGS for higher resolution RBC typing in the future.

## METHODS

### Study design

The design of this study included multiple interrelated components (Figure 1). We collated 2,796 WGS data from the ASPREE study, and 2504 from the 1KGP3 phase 3 (1KGP3) databases. Descriptions of the database ASPREE^35^ and 1KGP3^37^ are included in Supplementary Table 1. The gVCF and BAM files were used to predict genotype and phenotype using RBCeq. We collated 96 709 high-quality coding variants from genomes of ASPREE participants. Allele frequencies and clinical relevance of the variant were obtained using ANNOVAR^38^ (gnomAD genome collection [v2.1.1]). Additionally, sequencing coverage and sequence identity of the blood group genes were extracted from the alignment files using BAMSTAT39 and IGV40, respectively.

**Table 1:** The Frequency of null phenotypes identified in ASPREE data compared with and 1KG dataset.

| Blood Group System | Phenotype | RBCeq Prediction (%) |  |  |  |  |  |
| --- | --- | --- | --- | --- | --- | --- | --- |
|  |  | ASPREE<br>n = 2796 | AFR<br>n = 661 | AMR<br>n = 347 | EUR<br>n = 503 | EAS<br>n = 504 | SAS<br>n = 489 |
| CO | Co(a-b+) | 0.21 | - | - | - | - | - |
| YT | Yt(a-b+) | 0.14 | - | - | 0.20 | - | 0.20 |
| VEL | Vel- | 0.04 | - | - | - | - | - |

AFR: African, AMR: American, EAS: East Asian, EUR: European, SAS: South Asian

**Figure 1:**
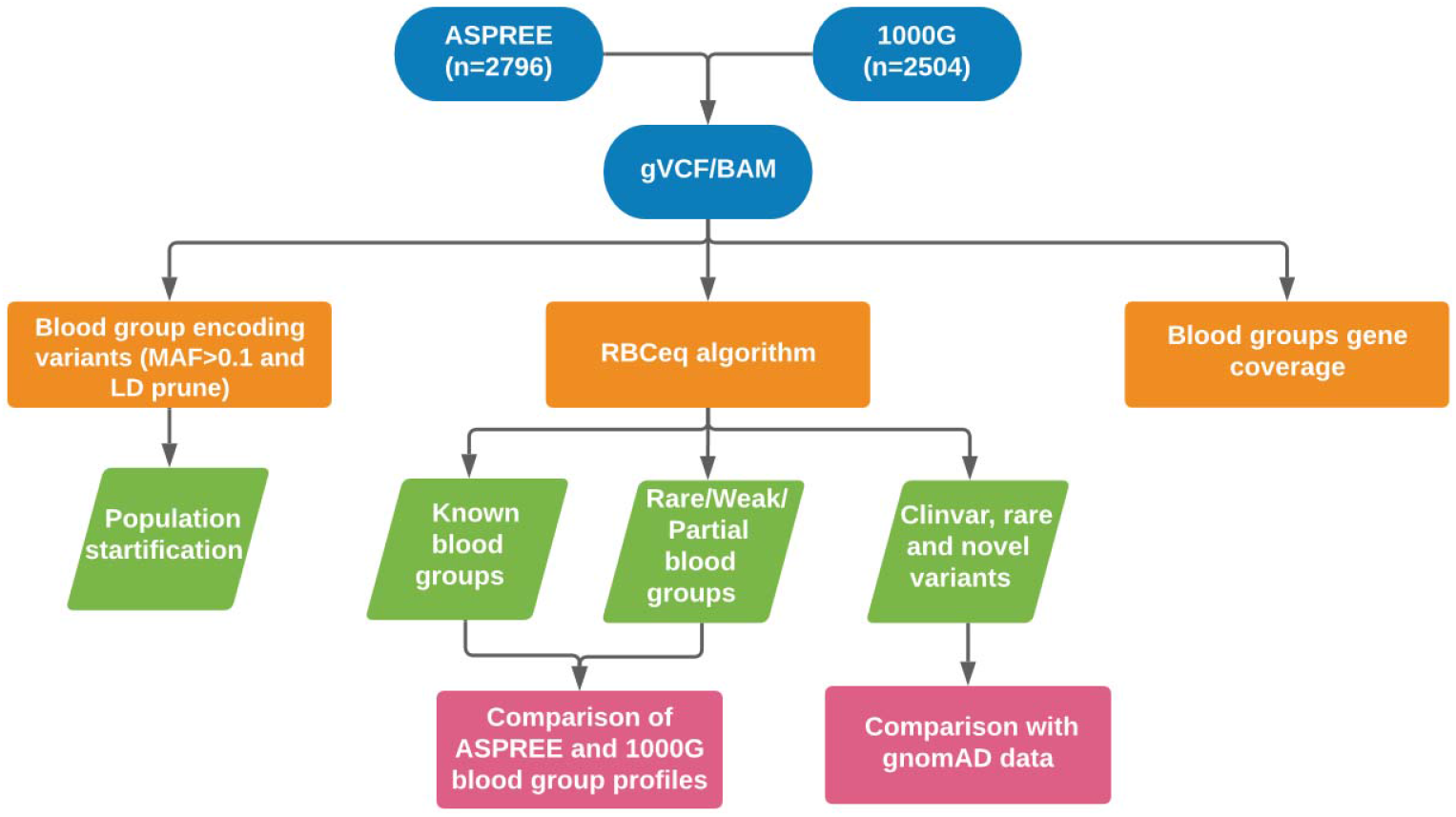
An overview of workflow to comprehensively characterise population specific blood group variants and phenotypes. The population stratification was performed using plink; blood group profiling using RBCeq and blood group gene coverage calculation using BAMstat and bedtools.

To evaluate the distinction between the ASPREE dataset and other global populations, we merged 789 linkage disequilibrium pruned coding variants of the ASPREE and 1KGP3 datasets, all of which had minor allele frequencies (MAF) ≥ 0.1. We then utilized PLINK v1.9^41^ to generate a principal component analysis (PCA) plot from the resultant merged data and plotted using ggplot2^42^ (Figure 2). Next, we created a Circos plot containing variant frequencies and the gene annotation using R libraries circlize^43^, tidyverse^44^, dplyr^45^, ComplexHeatmap^46^, and stringr^44^ (Figure 3). The prediction of RHD zygosity and C/c antigen was confirmed by RBCeq using the RH blood group gene coverage plot. Initially, all sample RH genes coverage was calculated using BedTools^47^ with bin size of 1. The coverage data was then smoothed and downscaled by averaging every 300 bases. The smoothed data were plotted using R libraries.

**Figure 2.**
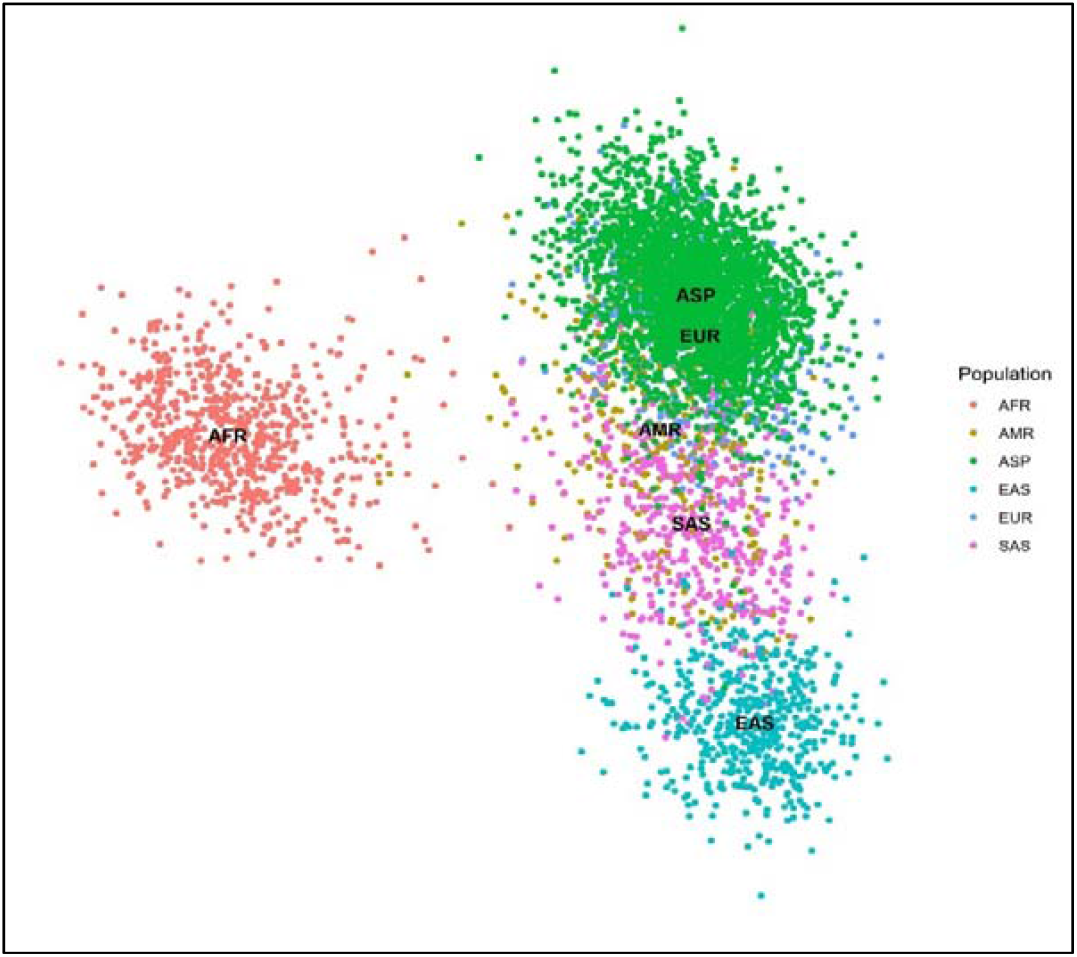
PCA plots showing 789 (MAF ≥ 0.1 and LD prune) markers in 5300 individuals from the ASPREE and 1KGP3 datasets. The X-axis denotes the value of PC1, while Y-axis denotes the value of PC2, with each dot in the figure representing one individual. The first two principal components shown here account for ∼80% of the observed variance in the combined dataset. (AFR: African, AMR: American, EAS: East Asian, EUR: European, SAS: South Asian, LD: linkage disequilibrium, MAF: minor allele frequency, PCA: principal component analysis.

**Figure 3:**
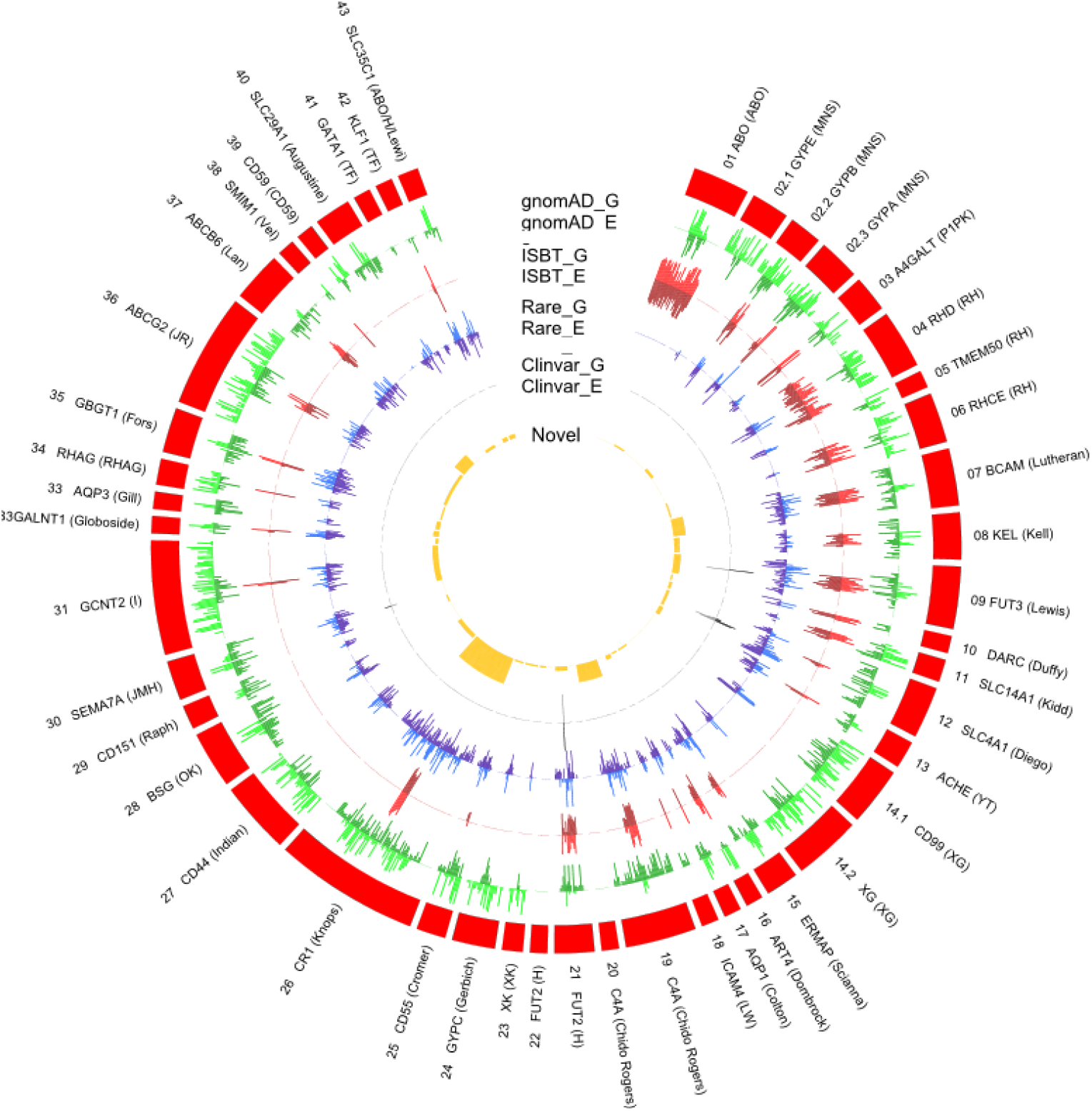
The distribution of gnomAD (genome and exome) datasets genetic variants and their frequency in RBC antigen encoding genes. The outer ring (red) represents the RBC antigen encoding genes; box length represents the number of variants observed; G denotes gnomAD genome frequency, and E denotes gnomAD exome frequency. The outer green (light/dark) circle indicates the distribution of variants frequency across different blood group genes from the gnomAD data. The red (light/dark) circle indicates the number of variants with ISBT associations relative to all gnomAD variants. The blue (light/dark) circle indicates the distribution of rare non-ISBT variants in all six populations. The dark grey circle indicates the number of non-ISBT variants annotated to the ClinVar database. The yellow circle shows the distribution of the number of novel variants. The data was visualised using Circos (reference)

### Genotyping using RBCeq

The RBCeq algorithm determined the genotype and predicted phenotype of the ASPREE and 1000 genome phase three cohort individuals(https://www.rbceq.org/). RBCeq predicts blood group profiles based on both SNV and CNV data. The gVCF of each sample was used as input along with the blood group gene coverage calculated using BAMTrimmer (https://github.com/MayurDivate/BamTrimmer). The RBCeq predicted known blood group profiles for 36 blood group systems and two transcription factors, and identified ClinVar, Rare (<=0.05 in gnomAD dataset) and deleterious novel variants.

## RESULTS

### The blood group gene variant landscape in healthy older Australians

The PCA plot revealed blood group genetics for the ASPREE and European participants from the 1KGP3 datasets exhibited a unique genomic identity distinct from other global population groups. The nearest correlation to ASPREE and European blood group genetics was found in the American population. Clear differentiation between the African, East Asian and South East Asian data was observed (Figure 2).

### Distribution of null phenotypes of high prevalence antigens

Caucasian people with rare null phenotypes are at risk of alloimmunisation if transfused. We found 0.21% ASPREE participants to express rare Colton Co(a-b+) phenotype. The rare Vel-negative blood type was found only among the ASPREE participants (0.04%).^48^ In the YT system, the Yt(a-b+) subtype was present in 0.14% of ASPREE, 0.20% of European, and 0.20% of South Asian participants. Transfusion of alloimmunised Co(a-) and Vel-individuals would require antigen negative blood to avoid HTR.

### Distribution of weak, partial and null antigens

Analysis of the distribution of the weak, partial and null antigens among the representative populations from the two databases showed a 93.34% distribution of the Duffy null phenotype Fy(a-b-) amongst African participants, which was higher than that reported in previous studies (68%-70%) (Table 2). Apart from African populations, Duffy null was only found in the American population (African-American). The Duffy null phenotype was not found amongst all other populations consistent with earlier reports.^11,49^ We report for the first time the distribution of a rare parabombay H+□ type (*FUT2*01W*.*02*.*01*), which was found only among the ASPREE (0.07%, n=2) and East Asian (19.05%, n=96) populations and was not represented in the European dataset. We also found a very low percentage of the extremely rare Kel 6,-7 antigen (0.15%) previously reported frequency in African populations is 19% and in Caucasians <1%^50^. The rare LU:-13 phenotype was found among the ASPREE participants (0.21%) previously reported only amongst European populations (0.40%).

**Table 2:** Comparing Weak, Partial and Null Blood group antigens distribution in the ASPREE and 1KG datasets.

| Blood group System | Phenotype | RBCeq Prediction |  |  |  |  |  |
| --- | --- | --- | --- | --- | --- | --- | --- |
|  |  | ASPREE<br>n = 2796 | AFR<br>n = 661 | AMR<br>n = 347 | EUR<br>n = 503 | EAS<br>n = 504 | SAS<br>n = 489 |
| LU | LU:-13 | 0.21 | - | - | 0.40 | - | - |
| H | H+□ | 0.07 | - | - | - | 19.05 | - |
| FY | Fy(a-b-)* | - | 93.34 | 1.73 | - | - | - |
| KEL | KEL6,-7 | - | 0.15 | - | - | - | - |
| JK | Jk(a+□) | 0.54 | 4.54 | 4.61 | 1.19 | 15.87 | 8.38 |
| JR | Jr(a+□) | 1.43 | - | 1.73 | 1.19 | 7.94 | 0.61 |
| ABO | A3 | - | 0.15 | - | - | - | - |
| ABO | B3 | 1.65 | 0.15 | - | - | - | - |

\*only for erythroid cells, AFR: African, AMR: American, EAS: East Asian, EUR: European, SAS: South Asia

Not a single individual with a very rare Jk(a–b–) phenotype, which is most frequent in Polynesian populations, was found among all six populations studied (Table 2). The Jk(a+wb-) phenotype which causes weak or partial expression of the Jk^a^ antigen, was represented at frequencies 0.54% (ASPREE), 4.54% (African), 4.61% (American), 1.19% (European), 15.87% (East Asians) and 8.38% (South Asian) within the six population datasets. We found the highest prevalence of the Jr(a+□) phenotype among East Asian populations (7.9%) while the African participants completely lacked the phenotype. Similar prevalence of Jr(a+) was observed among the other populations in our study (ASPREE [1.4%], European [1.2%], South Asian [0.6%]). ^11^ The A3 (0.15%) subtype was only observed in the African population, while the B3 phenotype was observed in both ASPREE (1.65%) and African (0.15%) populations.

### Prevalence of RHD/RHCE blood group phenotypes

We compared the frequency of Rh blood group phenotypes frequencies in ASPREE and 1KGP3 samples with previously reported data (Table 3). We analyzed the D gene percentage with respect to homozygous null expression within the six populations. Our results indicated that 99.8% of East Asian, 96.52% of African, 93.95% of American, and 94.48% of South Asian participants were homozygous for the RHD gene. Previous reports state approximately 85% of the Caucasian population is D-positive and RBCeq also predicted 83. 9% in European populations consistent with earlier reports (**Supplementary Figure 1**). In the 474 (16.92%) D-negative samples, 473 were due to deletion of the RHD gene and one sample was homozygous for the RHD*01N.20 allele. This is possibly the first to report for partial RHD antigen named DIIItype4 and DUC2 in Australian Caucasian cohorts. In Aspree two hemizygous calls for DIIItype4 and two homozygous calls for DUC2 were observed. Other than ASPREE, only in African populations DIIItype4 and only in American population DUC2 were observed. These two partial D types are rarely observed in Caucasians (0.1% European Americans), the same rarity we observed in other 1KG populations.

**Table 3:** RHD/RHCE blood group phenotype frequency prevalence in ASPREE and 1KGP3 samples compared with previously reported data *RHD*DIII* ()0.04% and *RHD*DUC2*

| Blood group gene | Predicted Phenotype | RBCeq Prediction |  |  |  |  |  | Previously Published |  |  |
| --- | --- | --- | --- | --- | --- | --- | --- | --- | --- | --- |
|  |  | ASPREE<br>n=2796 | AFR<br>n=661 | AMR<br>n=347 | EUR<br>n=503 | EAS<br>n=504 | SAS<br>n=489 | Caucasians | Africans | Asian |
| RHD | DD | 83.09 | 96.52 | 93.95 | 83.9 | 99.8 | 94.48 | 85.00 | 92.00 | 99.00 |
| RHD | D- | 16.92 | 3.48 | 6.05 | 16.10 | 0.20 | 5.52 | 15.00 | 8.00 | 1.00 |
| RHD | DIII type4 | 0.07 (Two heterozygous call) | 2.87 ( Four homozygous and 15 heterozygous) | - | - | - | - | - | - | - |
| RHD | DUC2 | 0.07 | - | 0.30 (Two heterozygous call) | - | - | - | - | - | - |
| RHCE | C+ | 32.98 | 23.30 | 56.48 | 39.36 | 91.07 | 60.53 | 68 | 27 | 93 |
| RHCE | c+ | 45.60 | 84.87 | 55.62 | 49.30 | 40.67 | 28.63 | 80 | 96 | 47 |
| RHCE | E ce | 25.04 | 13.16 | 10.09 | 24.25 | 1.19 | 9.41 | 10.00 | 4.00 | 30.00 |
| RHCE | C E c e | 26.18 | 17.39 | - | 28.43 | - | - | 10.00 | NA | NA |
| RHCE | C partial c<br>partial e | - | 0.61 | - | - | - | - | NA | NA | NA |
| RHCE | C partial<br>ec | - | 0.30 | - | - | - | - | NA | NA | NA |
| RHCE | Cw partial<br>C e MAR- | - | - | - | 0.20 | - | - | NA | 19.00 | 3.00 |
| RHCE | partial c<br>partial e | - | 4.68 | - | - | - | - | NA | NA | NA |
| RHCE | partial e c | - | 2.12 | - | - | - | - | NA | NA | NA |

NA: The population-level frequency was not available in the literature; AFR: African, AMR: American, EAS: East Asian, EUR: European, SAS: South Asian

The C antigen occurs when reads of exon 2 of RHCE are mapped to exon 2 of RHD. It results in an increase of aligned reads to RHD exon 2 with no alignment to RHCE exon 2 sequence51-53 (Supplementary Figure 2). Our data for C/c antigen distribution are similar to earlier reports except for the ASPREE, European and South Asian participants. Our predictions showed the prevalence of C/c antigen among the ASPREE and European participants was 40-50% lower than previous report. In South Asian, the C/c antigen prevalence was also 20-25% lower than previous reports. We found the prevalence of the Ece and CEce phenotypes was two-fold higher in the ASPREE and European populations when compared with previous data^54^. We also detected partial c and e phenotypes in the 1KGP3, only in African population. Two individuals from the European population exhibited the *RHCE*02*.*08*.*01* allele at a homozygous level which encodes low prevalence antigen Cw, a partial C and MAR null phenotype. *RHCE*02*.*08*.*01* is clinically significant and is associated with HDFN, has been reported more frequently in the Polish population. There is a demand for MAR-blood in Poland ^55^. The frequency of *RHCE*02*.*08*.*01* allele phenotype has not been previously reported in a large scale study.

### Comprehensive analysis of MNS phenotype

Table 4 shows the frequency of phenotype M+N-S+s- and M+N+S+s- in additional ethnic (EAS/SAS/EUR/AMR) data. The frequency of M+N-S+s+, M+N+S-s+, and M+N+S+s+ was previously reported for Caucasian and African population; we observed a two-fold lower prevalence of all three blood types in European participants compared with earlier reports (4.97% vs 14.0%, 7.75% vs 22.0%, and 12.92% vs 24.0% respectively). Conversely, the prevalence of M-N+S+s+ phenotype was found to be five-fold higher in the European participants (30.02%) in our study compared with previous data (6.0%). We found prevalence of all four MNS phenotypes in the ASPREE participants to be similar to those reported for Caucasian populations earlier.

**Table 4:** Comparison of MNS blood group phenotypes and frequencies observed in the ASPREE and 1KGP3 datasets.

| Blood group | Phenotype | RBCeq Prediction |  |  |  |  |  | Previously Published [where? REF) |  |  |
| --- | --- | --- | --- | --- | --- | --- | --- | --- | --- | --- |
|  |  | ASPREE<br>n=2796 | AFR<br>n=661 | AMR<br>n=347 | EUR<br>n=503 | EAS<br>n=504 | SAS<br>n=489 | Caucasians | Africans | Asian |
| MNS | M+N-S+s- | 5.90 | 0.15 | 2.31 | 1.79 | - | 2.25 | 6 | 2 | NA |
| MNS | M+N+S+s- | 3.11 | 0.45 | 4.32 | 3.38 | - | 4.50 | 4 | 2 | NA |
| MNS | M+N-S+s+ | 14.70 | 1.21 | 8.07 | 4.97 | 1.19 | 5.11 | 14.00 | 7.00 | NA |
| MNS | M+N+S+s+ | 23.86 | 4.54 | 13.26 | 12.92 | 1.98 | 11.45 | 24.00 | 13.00 | NA |
| MNS | M+N+S-s+ | 23.39 | 11.04 | 6.63 | 7.75 | 18.06 | 12.07 | 22.00 | 33.00 | NA |
| MNS | M-N+S+s+ | 5.04 | 20.12 | 22.19 | 30.02 | 3.57 | 25.15 | 6.00 | 5.00 | NA |

NA: The population-level frequency was not available in the literature; AFR: African, AMR: American, EAS: East Asian, EUR: European, SAS: South Asian

### Investigation of novel and clinically significant blood group variants amongst healthy older Australians

We characterized variants within the ASPREE population dataset that have not previously been reported to be associated with blood group alleles but have the potential to affect the formation of antigenic structures (Figure 3). We detected nine variants that had clinical associations (Supplementary Table 2). There were 688 rare variants with frequencies of ≤ 0.05 (genomAD) amongst the ASPREE participants’ data (Supplementary Table 3). Notably, we identified 149 novel variants that were computationally predicted to be deleterious (Figure 3, Supplementary Table 4). Predictably, no clinically significant, rare and novel variants were found to be associated with the genetically complex and most studied ABO blood group system, and only one novel and 16 rare variants were associated with the RH blood group system, which is also amongst the most studied blood group systems.

## Discussion

In this study, we compared the blood group genotype profile of Australian participants from the ASPREE study with African, American, Asian, and European population data from the 1KGP3 and gnomAD databases. The population stratification analysis revealed the genetic makeup of the ASPREE and European participants distinct from the other populations and its likely because many ASPREE participants were of European descent (Figure 2). We also observed a clear differentiation between the African participants and other populations, consistent with previous reports on the evolution of human blood group genetics.

We analyzed the distribution of rare phenotypes amongst the six population datasets (Table 1). We found the Colton Co(a-b+) phenotype was high frequency amongst the ASPREE participants, consistent with European ancestry. The Co(a-b+) phenotype is extremely rare and we found this predicted phenotype dataset at a frequency of 0.21% (n=6) in the ASPREE dataset, also consistent with frequencies reported in Europe. Previously, seven separate studies on 13 460 donors from Northern Europe and North America showed 91.3% prevalence of Co(a+b-) and 8.5% prevalence of Co(a+b+) and only 0.2% prevalence of Co(a-b+).^56^ A study on 1706 African Americans showed a 100% prevalence of the Co(a+) phenotype.^56^.

In ASPREE, participants with the rare Vel-negative phenotype was also predicted. Antibodies to the Vel antigen are known to be associated with complement activation. Vel is a high frequency antigen, inherited as an autosomal recessive trait, that shows variable strength, ranging from strong to weak.^21,57^ Vel-negative individuals can develop anti-Vel antibodies after transfusion or pregnancy and may develop acute HTRs if transfused with Vel-positive blood.^21,57^ Therefore it is of value to screen the population to identify Vel-blood donors and add to the national frozen blood bank to fulfil future requests.

We also found the low prevalence ABO alleles A3 and B3 only among the African (A3:0.15%, B3:0.15%)and ASPREE (B3:1.65%) participants. The B3 prediction of ASPREE is with B genotype possibilities, and may require further serology work to confirm the genotype reflects phenotype. Earlier studies have indicated that distinct ABO variants were associated with different populations.^58^

It is important to type clinically significant RBC antigens of patients and blood donors in a pretransfusion setting to ensure blood components are matched appropriately for transfusion, to prevent transfusion related adverse events and reduce alloimmunisation.^19^ We typed clinically significant rare and weak blood group variants among the six populations (Table 2). The RBCeq prediction showed a significantly higher (93.34%) prevalence of the Duffy null Fy(a-b-) phenotype in the African dataset compared with earlier reports (68%-70%)^11^.

The observed Fy(a-b-) phenotype was predicted from genotype FY*02N.01/FY*02 by RBCeq. The phenotype FY(a-b-) sometimes may be genotypically FY*01N.01/*02, in such cases the patient is unlikely to form anti-Fy3 as the patient will express Fy^b^ in tissues. We found para-Bombay phenotypes (H+□) amongst the ASPREE (0.07%) and East Asian (19.05%) participants. However, in the Chinese population, a greater frequency of para-Bombay than Bombay has been reported.^11,59^ Our results showed a very low percentage of the extremely rare KEL6,-7 antigen only amongst the African dataset (0.15%).

We also identified the extremely rare LU:-13 phenotype among the ASPREE participants (0.21%) which has only been observed previously among European populations. The Lutheran blood group system comprises 20 antigens defined by largely clinically benign antibodies. However, no specific data is available on the adverse events of transfusion of anti-LU13 due to the extreme rarity of the antibody. Therefore, there is a recommendation to transfuse Lu(a-b-) blood and do family studies to identify potential donors if a patient/donor is found to lack the LU:13 antigen.

We didn’t find any evidence of the Jk(a–b–) phenotype, which is found in Polynesians, amongst the six populations studied. Red Cross Lifeblood has appealed for blood donors of Polynesian heritage to donate in an attempt to identify more Jk(a-b-) donors as there is demand for blood of this phenotype. The weak or partial expression, Jk(a+^w^) antigen was observed in all six populations with the highest prevalence seen among the East Asian participants. The Jk(a+^w^) phenotype can be a basis of discordance between serological and molecular methods. A phenotype with weakened Jk^a^ expression may constitute a risk for HTRs if antigen-positive units are not identified during pre-transfusion/donation serology tests.

Anti-Jr^a^ has been reported to cause fatal cases of HDN and delayed HTRs^60,61^. We found the highest prevalence of the Jr(a+□) phenotype among the East Asian (7.9%) populations while the African population completely lacked the phenotype. Similar prevalence of prediction of the Jr^a^ antigen was observed among the other populations in our study. As with previous studies, we found the Kel6,-7phenotype only in the African dataset.^11^

Fetal anemia is one of the most serious consequences of paternal blood group incompatibility with the mother. The most clinically significant antibodies involved in HDFN are the RhD antigen, and other minor Rh antigens C, E, c, e antigens. Antigens of other blood group systems (Duffy, Kidd, Kell, M and S) rarely cause significant problems, however it is a requirement^9^ to test and monitor pregnant women for the formation of these antibodies, as some can cause HDN.^62^. For anti-D, anti-K, and anti-c, there is a >50% risk of mild to severe HDFN developing in case the fetus inherits the target paternal red cell antigen. We analyzed the RHD gene haplotypes percentage in all six populations (Table 3). Our results indicated the majority (99. 8%) of the East Asian participants, and most of the African (96.52%), American (93.95%) and South Asian (94.48%) participants were homozygous for the *RHD* gene. However, the European (83.9%) and ASPREE (83.09%) participants had lower homozygous *RHD* gene frequencies. Genetic variants that result in weakened or partial antigen expression are problematic in transfusion medicine and can result in mistyping of RBCs. We also identified rare partial RHD types *DUC2* (0.07%) and DIIItype4 (0.07%), observed in Caucasians (approx.0.1% European Americans). Partial D proteins lack certain D epitopes which need to be characterised in order to guide the selection of blood for transfusion and in antenatal care, as if these individuals are transfused with D+ blood (all D epitopes present) they can be alloimmunised to the epitope they lack. And individuals with these antigens should ideally receive D-negative donor RBCs to avoid alloimmunization. In some cases. individuals with weak D should receive D-negative RBCs to avoid alloimmunisation but are often mistyped as D positive with a risk of producing an alloanti-D antibody.^25,53^ Additionally, It is important to differentiate weak D from weak D/Partial and Partial D types in pregnant women to provide anti-D prophylaxis during pregnancy and in women of childbearing potential to prevent D alloimmunisation..^25^

RBCeq predictions for the C-antigen prevalence among the six population datasets followed similar patterns to earlier studies for the predominantly African and East Asian (Table 3). However, C-antigen prevalence in our predictions for the ASPREE, European and South Asian datasets was significantly lower than previously reported values^63^.

The points of contrast presented for the blood group profiles of the ASPREE cohort are of potential clinical significance. In our study, the ASPREE and European participants had two-fold higher prevalence of C-E+c+e+ and C+E+c+e+ compared with previous reports for Caucasian populations. We could also found two instances of Cw, a partial C and MAR null within the European population never reported before using genomics data.

To the best of our knowledge, this is the first study describing the frequency of M+N-S+s- and M+N+S+s- in different ethnic backgrounds using genomic data (Table 4). We found the prevalence of all MNS phenotypes among the ASPREE participants to be similar to those reported earlier for Caucasian populations.

The prevalence of blood group antigen variants based on exon and splice-site variants were compared to global population groups (Figure 3). The results reflected a high level of conservation in the majority of blood group systems. We detected nine variants that had pathogenic clinical associations (Supplementary table2). We also detected 688 rare variants (238 with greater than two-fold difference from the gnomAD exome) with minimum allele frequencies of ≤ 0.05 in any of the five populations (Supplementary Table 3). Most importantly, we identified 149 variants that were computationally predicted to be novel and deleterious (Supplementary Table 4). Predictably, no clinically significant, rare, and novel variants were found to be associated with the most studied genetically complex ABO blood group system, and only one novel and 16 rare variants was associated with the RH blood group system. This observation is expected from these two most commonly studied systems.

While this is possibly the first comprehensive multi-ethnic study where genetic data from six different populations was compared, certain limitations need to be acknowledged. The ASPREE dataset, though heterogeneous, predominantly included participants of European descent with Caucasian population values. The dataset also did not include investigation of the hybrid RH and MNS changes. Serological testing and exon level detections were not performed to back up the RBCeq phenotype predictions. However, despite the limitations, our results are providing new insights into blood group genetics and antigen frequency.

The frequency of inherited disorders like SCD and thalassaemia are increasing in Australia due to increased migration between the populations.^64^ RH alloimmunisation is not completely prevented in patients with SCD although matched for extended blood groups due to antigen polymorphism in the RH system. These are examples of the significant differences between Australia’s blood donors, who are mostly Caucasian and African SCD transfusion recipients.^64^ Alpha thalassaemia has been identified in Australian patients in Aboriginal and Torres Strait Islander communities in the Northern Territory and northern Western Australia.^65^.^65^ Rare blood group identification will improve rare blood unit management. Since language and culture are very diverse in Australian communities, additional research is needed to guide transfusion practice and improve transfusion outcomes in Australia.

## Data Availability

The Medical Genome Reference Bank

## Notes

### Competing Interest Statement

The authors have declared no competing interest.

### Funding Statement

1) NSW Office of Health and Medical Research - Sydney Genomics Collaborative grant (2014).
2) National Institute on Aging and the National Cancer Institute at the National Institutes of Health (grant number U01AG029824);
3) The National Health and Medical Research Council of Australia (grant numbers 334047, 1127060)
4) Advance Queensland Research Fellowships

### Author Declarations

IRB00002519; ethics #2006/745MC The study was approved by the Alfred Hospital Research Ethics Committee (Project ID: 390/15) and registered on Clinicaltrials.gov 56 (NCT01038583). Participants provided biospecimens and written informed consent for genetic analysis.

